# Protocol for the AutoRayValid-RBfracture Study: Evaluating the efficacy of an AI fracture detection system

**DOI:** 10.1101/2023.08.15.23294116

**Authors:** Huib Ruitenbeek, Liv Egnell, Katharina Ziegeler, Mathias Willadsen Brejnebøl, Janus Uhd Nybing, Anders Lensskjold, Pavel Klastrup Lisouski, Michael Lundemann, Kay Geert A. Hermann, Mikael Boesen, Edwin H.G. Oei, Jacob J. Visser

**Author notes:** Corresponding author Huib Ruitenbeek, Department of Radiology & Nuclear Medicine, Erasmus Medical Center, Rotterdam, The Netherlands. **Administrative information**. **Funding** This project has received funding from the European Union’s Horizon 2020 research and innovation programme under grant agreement No 954221 for the EIC SME Instrument project AutoRay. The work only reflects the authors’ view and the European Commission is not responsible for any use that may be made from the information it contains. **Roles and responsibilities** The study is a collaboration between Radiobotics ApS (Sponsor) and the following three European clinical sites; Department of Radiology, Bispebjerg and Frederiksberg Hospital, Copenhagen, Denmark (BFH) Department of Radiology, Charité - Universitätsmedizin Berlin, Berlin, Germany (CUB) Department of Radiology & Nuclear Medicine, and Erasmus Medical Center, Rotterdam, The Netherlands (EMC). Sponsor Contact information Trial Sponsor: Radiobotics ApS, Contact name: Michael Lundemann, Address: Esplanaden 8C, 1263 Copenhagen K, Denmark.

## Abstract

**Background:** Rapidly diagnosing fractures in appendicular skeletons is vital in the ED, where junior physicians often interpret initial radiographs. However, missed fractures remain a concern, prompting AI-assisted detection exploration. Yet, existing studies lack clinical context. We propose a multi-center retrospective study evaluating the AI aid RBfracture™ v.1, aiming to assess AI’s impact on diagnostic thinking by analyzing consecutive cases with clinical data, providing insights into fracture detection and clinical decision-making.

**Objectives:** To provide new insights on the potential value of AI tools across borders and different healthcare systems. We will evaluate the performance of the AI aid to detect fractures on conventional x-ray images and how its use could affect handling of these cases in a healthcare setting. In order to explore if the use of a trained and certified AI tool on clinical data exposes new challenges, a daily practice clinical scenario will be approached by minimising selection criteria and using consecutive cases. A multicenter, retrospective, diagnostic accuracy cross-sectional design incorporates clinical context.

**Methods:** The multicenter study spans three European sites without onsite hardware. AI system RBfracture™ v.1 maintains consistent sensitivity and specificity thresholds. Eligibility involves age ≥21 with x-ray indications for appendicular fractures. Exclusions include casts, follow-up x-rays, nearby hardware. AI aids retrospective fracture detection. Reader sessions include radiology and emergency care residents and trainees reading with and without AI. Fractures are marked, rated, with expert-established reference standards.

**Data:** Sequential patient studies at three sites yield 500 cases per site. Data includes anatomy, referral notes, radiology reports, and radiographic images. Expert readers use annotations, clinical context for standards. Statistical methods include dichotomized confidence ratings, sensitivity, specificity calculations, site-based analysis and subgroup considerations.

**Reference Standard:** Two experienced readers annotate fractures; if their annotations overlap by 25% or more, the common area is the reference. Discrepancies are resolved by a local expert. Individual fractures are labelled.

## Introduction

### Background and rationale

Fractures of the appendicular skeleton are typically diagnosed in the emergency department (ED). In the typical ED setting, junior physicians (in training) from a variety of clinical disciplines (eg orthopaedic surgery, general surgery and emergency medicine) or radiology residents are responsible for the initial interpretation of radiographs, and subsequent urgent decision-making. Mattijssen-Horstink et al^1^ reported that from a total of 25,957 fractures treated in the ED in the Netherlands, 289 fractures (1.1%) were missed by ED treating physicians. These fractures were diagnosed later during radiologic reading by radiologists. Moreover, 49% of all missed fractures took place between 4 PM and 9 PM.^1^ Therefore, an accurate and efficient assistant technology in fracture detection is of interest.^2^

The use of AI for automated fracture detection might reduce the number of missed fractures in the clinic by acting as an aid/second reader and bringing attention to detected fractures. Two recent papers published in Radiology reporting on reader performance involving AI fracture detection software aim to describe this effect on fracture detection. The first paper, by Guermazi et al^3^, describes a retrospective study of 480 patients. Readers were presented the whole validation data set, with and without AI assistance, with a one-month minimum washout period. The authors conclude a 10.4% improvement of fracture detection sensitivity without specificity reduction.^3^ The second paper, by Duron et al^4^, describes a retrospective study of 600 patients. The sensitivity of emergency physicians improved from 61.3% to 74.3% with AI, an increase of 13%, and the sensitivity of radiologists improved by 4.3% from 80.2% to 84.6%.

In both papers, the study designs differ from the context in the clinical setting in two main ways. First, a stratified dataset was used to balance it based on prevalence and anatomic locations. This means the sample is not representative for the general patient population, making it difficult to infer the results to a real-world clinical setting. Second, the readers were not presented with any clinical data at time of reading. This is likely to cause underestimation of the clinicians performance which as a consequence could lead to overestimation of reader improvement with AI aid.

In this evaluation of an AI aid, we propose a retrospective study design that aims to address these two limitations. The AI aid under investigation, RBfracture™ v.1, is a medical software device that automatically analyses radiographs for fractures. If a fracture is detected the software outputs a report stating that a fracture has been detected and marks the location of fracture with a rectangle in an overlay (Figure 1).

**Figure 1.**
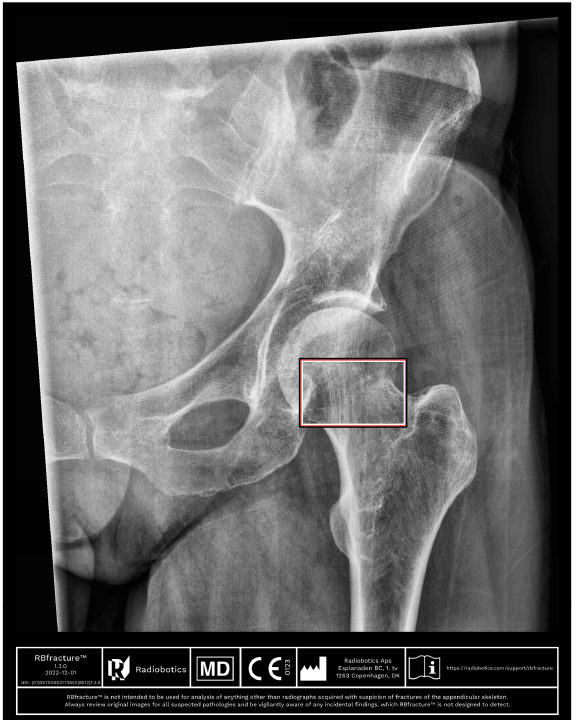
The AI aid under investigation is a medical software device that automatically analyses radiographs of the appendicular skeleton for fractures. If a fracture is detected in the image, the software outputs a report stating that a fracture has been detected and marks the location of fracture with a rectangle in an overlay for each radiograph where a fracture has been detected.

We propose an approach to assess the *Diagnostic thinking efficacy* or the *added value to the diagnosis*, as described by Fryback at al.^5^ In diagnostic imaging, this level of efficacy focuses on “whether the information produces change in the referring physician’s diagnostic thinking”^5^, which for applications of AI in radiology can be described as “reader performance with/without AI, or proving change in radiological judgement”.^6^ To fully evaluate the change in the referring physician’s diagnostic thinking, the readers will be presented with consecutive cases, including clinical data. Furthermore, we will explore how the introduction of AI influences the diagnostic thinking efficacy in terms of strengthening an existing hypothesis or reassuring the physicians in their diagnostic decisions^5^. Finally, this study is multi-country, extending the generalizability even more compared to current literature.

### Objectives

The aim of this inter-center reader study is to provide new insights on the potential value of AI tools across borders and different healthcare systems. In this study we will evaluate the performance of the AI aid to detect fractures on conventional x-ray images and to evaluate how its use could affect handling of these cases in a healthcare setting. In order to explore if the use of a trained and certified AI tool on clinical data exposes new challenges, a daily practice clinical scenario will be approached by minimising selection criteria and using consecutive cases. In contrast to previously published algorithm validation studies in the domain, where readers are blinded to clinical data^3,4^, this study mimics a scenario closer to a real clinical setting by providing the readers with the clinical referral notes along with the radiographs.

The study will be set up across three hospitals in three different European countries to explore if this has influence on the efficacy of the algorithm. The objectives are threefold:

1. To assess how decision support from the AI aid affects diagnostic thinking efficacy of radiologists and ED physicians.
2. To assess the accuracy of the algorithm to point out the reported fracture.
3. To compare algorithm performance in different health care systems and review generalizability.

#### Primary research question

- Does decision support from the AI aid increase diagnostic test accuracy of radiologists and ED physicians?

#### Secondary research question

- How does the performance of the algorithm and diagnostic thinking efficacy differ across the different healthcare systems?
  - To what extent is the AI aid able to detect reported fractures on plain radiographs in a daily practice clinical setting
- How accurate is the algorithm in annotating the same fractures as the readers?

### Study design

For this multicenter reader study, a retrospective, diagnostic accuracy cross-sectional design will be used. The readers will be presented with consecutive cases, together with clinical information from the referral request.

Additionally this study is multi-country, enlarging the generalizability even more compared to current literature (Figure 2).

**Figure 2.**
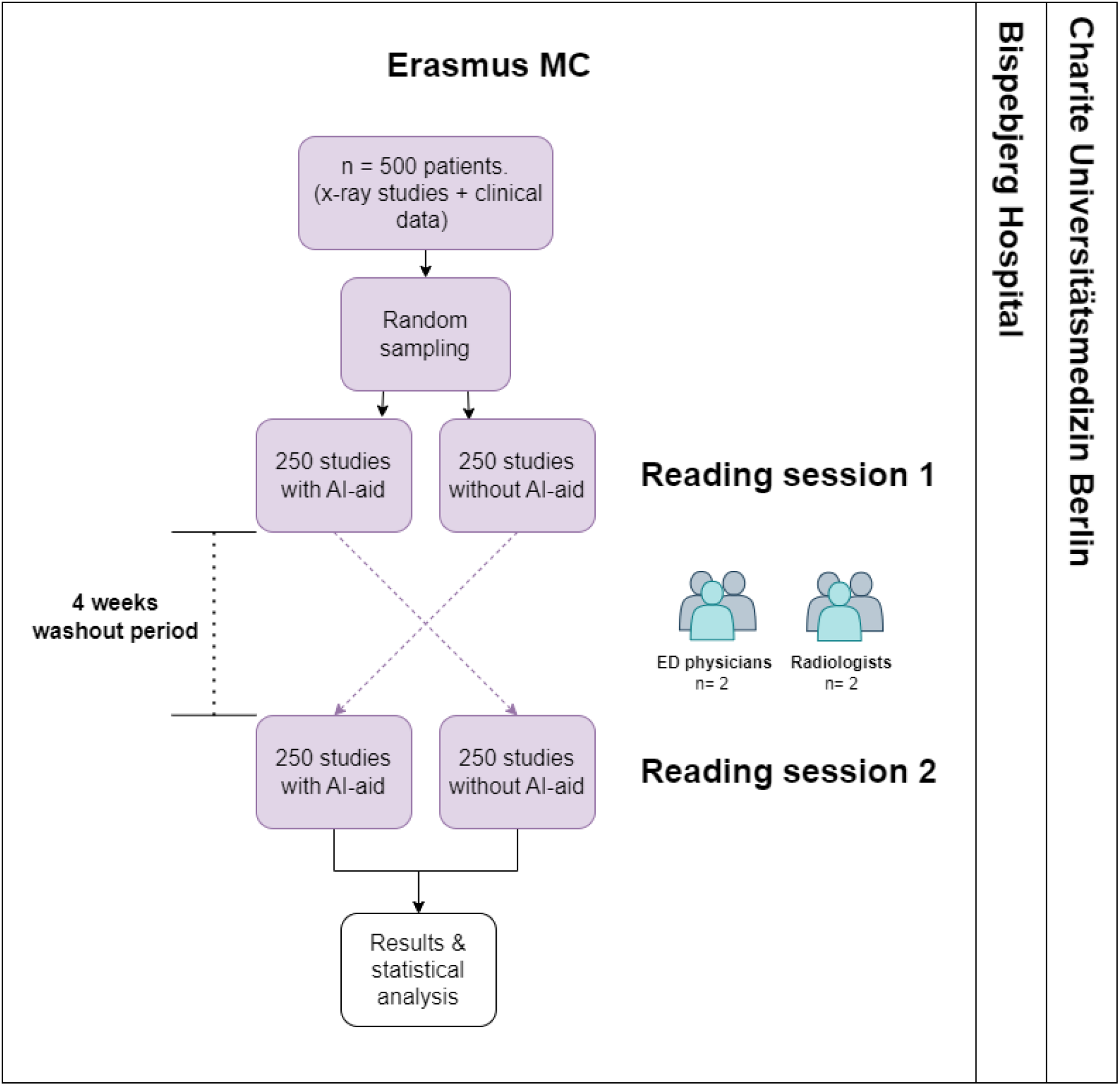
Schematic overview of the study design.

## Methods: Participants, interventions, and outcomes

### Study setting

This multicenter study will be carried out in the three European clinical sites listed in Section <u>Roles and responsibilities</u>.

There is no hardware installation required on site for this study. Since no site specific training or optimization of the threshold determining sensitivity and specificity of the AI system will be applied, this will allow for comparison of performance between the three healthcare systems.

### Eligibility criteria on the participant level

#### Inclusion

- Age: 21 years or above
- Indication of x-ray on suspicion of traumatic fracture (ED) in the appendicular skeleton and/or pelvis (including shoulder, upper arm, forearm, elbow, wrist, hand, finger, hip, pelvis, upper leg, lower leg, knee, ankle, foot and toe)

#### Exclusion

- Patients with a cast and/or patients referred for follow-up x-ray
- Hardware (alloplasties, screws, plates, nail, k-wires, pins etc.) present in close proximity to fracture

### Eligibility criteria on the data level

#### Inclusion

- Projections of the anatomy subgroup suspected for fracture.
- If images from multiple anatomical subgroups (as defined in Table 1) exist from the same patient, only images from one of these subgroups will be included. The selection of anatomical subgroups will be based on random sampling.

#### Exclusion

- A previous examination of the patient has already been included
- Hardware (alloplasties, screws, plates, nail k-wires, pins etc.) present in close proximity to fracture
- Poor radiographic quality, image clinically unsuitable. This can include patient positioning errors, inappropriate selection of technical exposure factors, patient motion, presence of artefacts, improper collimation of the radiographic beam, and absence of permanent anatomical side markers
- Patient exams containing fractures in anatomies outside of the intended use present in the study (eg rib, spine)

### Interventions

The AI aid under investigation, RBfracture™ v.1, is a medical software device that automatically analyses radiographs of the appendicular skeleton for fractures to aid health care practitioners in detecting fractures. This study uses retrospective data; hence no patient will undergo a supplementary examination or radiation exposure, nor will it influence their treatment. The reader participants will only interact with the produced output and should be suitably trained as specified under Section <u>Reader election</u>.

**Table 1.**
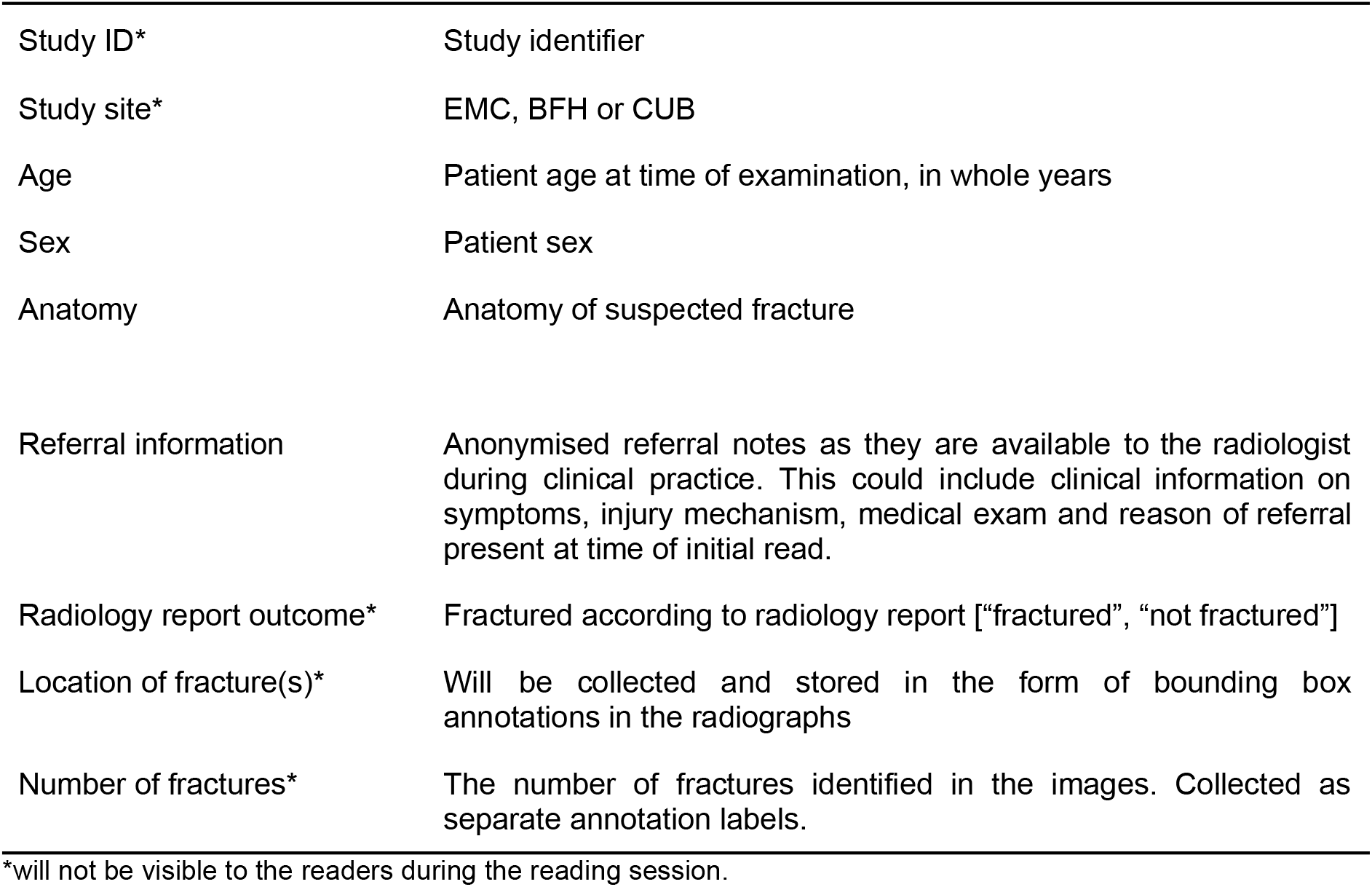
Overview of all information that is collected per case.

|  |  |
| --- | --- |
| Study ID* | Study identifier |
| Study site* | EMC, BFH or CUB |
| Age | Patient age at time of examination, in whole years |
| Sex | Patient sex |
| Anatomy | Anatomy of suspected fracture |
| Referral information | Anonymised referral notes as they are available to the radiologist during clinical practice. This could include clinical information on symptoms, injury mechanism, medical exam and reason of referral present at time of initial read. |
| Radiology report outcome* | Fractured according to radiology report ["fractured", "not fractured"] |
| Location of fracture(s)* | Will be collected and stored in the form of bounding box annotations in the radiographs |
| Number of fractures* | The number of fractures identified in the images. Collected as separate annotation labels. |

\*will not be visible to the readers during the reading session.

The intervention is using the AI software as an aid for fracture detection. The readers will participate in two reading sessions, separated in time by a washout period of minimum four weeks. In both sessions, all radiographs will be presented so that each radiograph will be read twice, once with and once without the AI aid. Randomisation to the intervention will be applied (based on computer generated random numbers) so that at the first read, the aid will be present for half of the patient studies and vice versa. Randomisation will be performed by a suitably trained person, performing the data management.

#### Reader annotations

For each patient, the reader participants will be presented with the radiographs together with the referral notes in a digital annotation platform. For each projection, the readers are asked to mark any detected fractures by placing a “dot” on the fracture line. The location and number of fractures is measured hereby.

For each fracture, the readers are also asked to rate their confidence in a four-level categorical scale; “definitely fractured”, “likely fractured, needs further imaging”, “likely not fractured, needs further imaging”, or “definitely not fractured”.^7^

##### Reader election

Four reader participants will be recruited at each site with the following profiles:

- One experienced musculoskeletal radiologist
- One radiologist in training
- One senior physician employed at the emergency department
- One junior physician employed at the emergency department

#### Reference standard

The reference standard will be collected as bounding boxes in the radiographic projections. Supporting clinical data, such as the radiology report from the initial visit and potential follow up imaging, and potentially also treatment data (if accessible under IRB approval), will be available to the reference readers as a support during the reference readings (see Figure 5). In addition, follow up images will be made available to the reference readers so they can access images if necessary. Since clinical routine can vary between sites, the specific time frame for the inclusion of follow up diagnostic images should be determined at each site. All images will be annotated by two reference readers, experienced in clinical MSK and/or trauma radiology, of which at least one will be recruited from the local radiology department.

The radiology report(s) will be available to the reference readers as a support during the reference readings. If the bounding boxes by the two reference readers overlap with an intersection over union of 25% or higher, the reference standard will be defined as the rectangle of the smallest area that encloses both of them (see Figure 3). Any discrepancies between these two are adjudicated by an experienced local reference reader to form the reference standard (Figure 4). Anyone who participates in the process of establishing the reference standard will be disqualified to participate as a reader in the reader sessions.

**Figure 3.**
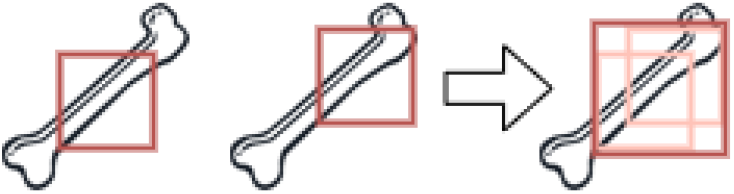
If the bounding boxes by the two reference readers overlap with an intersection over union of 25% or higher, the reference standard will be defined as the rectangle of the smallest area that encloses both of them.

**Figure 4.**
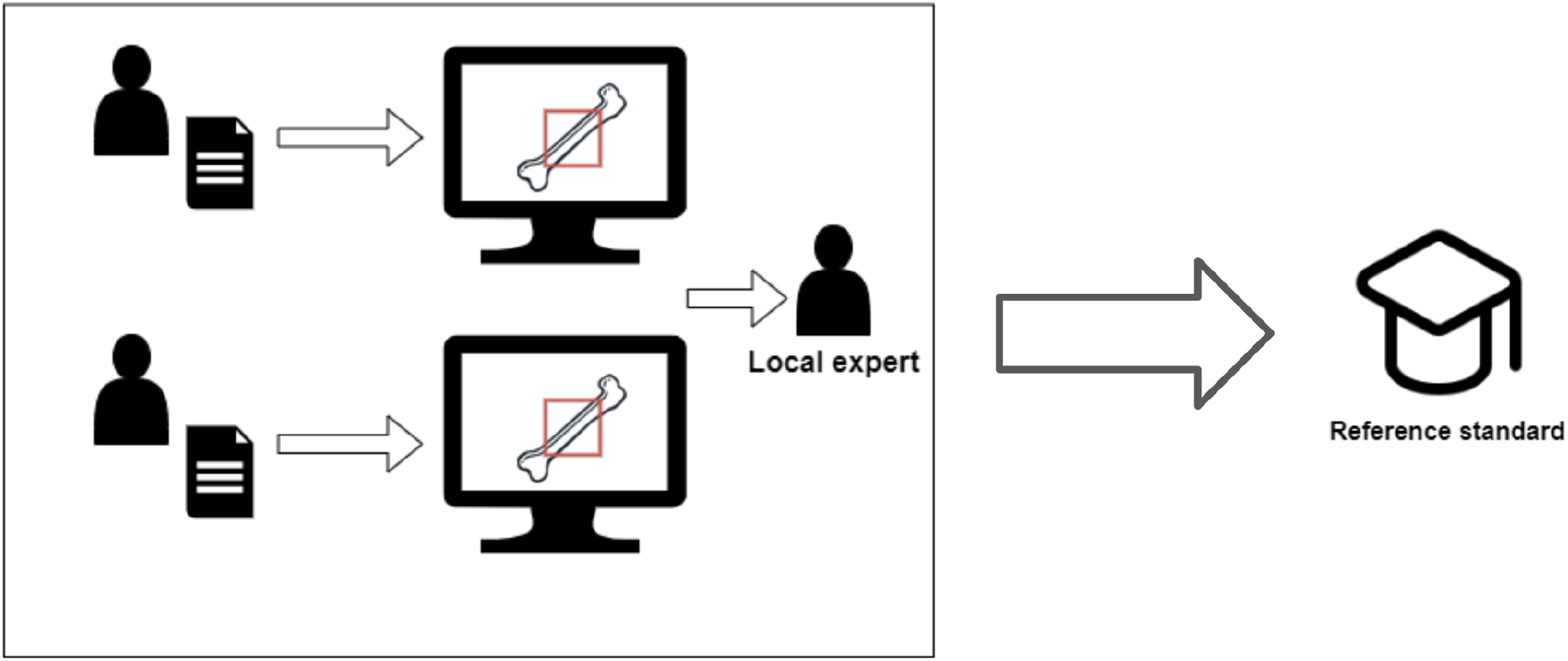
Two readers will annotate the fractures in all images to establish the standard reference. Any discrepancies will be adjudicated by a third reader.

All acute, subacute and healing fractures that are visible should be classified as positive and annotated in all projections to the extent that this is feasible based on the visibility. Chronic fractures or completely healed fractures that do not need clinical attention are not annotated. Individual fractures should be distinguished by labelling, “Fracture1”, “Fracture2”, etc, in order to allow for counting of the individual fractures that are detected by the readers.

If neither the supporting data, nor the consultation by experts provide a clear conclusion, the patient will be excluded.

All imaging data is derived from clinical production and therefore is expected to meet the clinical quality standard. In the process of defining the reference standard images with inadequate quality can be excluded.

#### Potential impact on decision making or clinical procedures

The AI device under intervention is designed to assist its users (eg radiologists and emergency physicians) in the clinical decision making for fracture detection. The detection of one or multiple fractures can be faster or the amount of fractures detected might change. There is no human-AI interaction that has influence on the AI result. Human-AI interaction is present during the reading of the x-ray image where the physician inspects both the AI output and the original x-ray to state a final diagnosis.

### Outcomes

The primary endpoint is the impact on diagnostic thinking. Specifically, this will be measured by the change in readers’ diagnostic performance (sensitivity and specificity) with and without decision support of the algorithm. In addition, the area under the receiver operating characteristics curve (AUC) of the readers will be calculated.

### Sample size considerations

We consider an estimated reading time of 3-4 cases per minute. This would result in a workload of 120 - 160 minutes to read all images per session (with or without AI aid) in the reader study. This was agreed upon by all contributing sites to be the maximum acceptable contribution. The true estimates of the parameters affecting the sample size and statistical power of the tests are not yet known, however with an estimated reader sensitivity of around 60-70% and specificity of 90-92%^3,4^ and an assumed prevalence of around 30%, a sample size of 500 is deemed sufficiently large to evaluate the primary endpoints, sensitivity and specificity. This sample size is also likely to provide sufficient statistical power for the secondary anatomical subgroup analysis, at least for the most commonly fractured anatomies.^8^

## Methods: Data collection, management, and analysis

### Data collection methods

At each site, studies of eligible patients will be included consecutively from the PACS until the target sample size of 500 patients is reached. The data will be sampled using inclusion and exclusion criteria described in Section <u>Eligibility criteria</u> by a suitably trained person and the study date where consecutive inclusion began and ended will be noted. The number of radiographic projections can vary because images are collected from multiple institutions with their own image acquisition protocols. All projections in the study of the selected anatomical subgroup will be included to reflect the real clinical setting where usually all projections would be available. If one patient has multiple eligible x-ray exams, one of the anatomy groups is included using random selection.

After image extraction, all cases are supplemented with the data listed in Table 1. The data will be grouped into six anatomical subgroups based on the imaged anatomy in order to avoid issues related to small sample sizes. The anatomical groups are listed in Table 2.

**Table 2:**
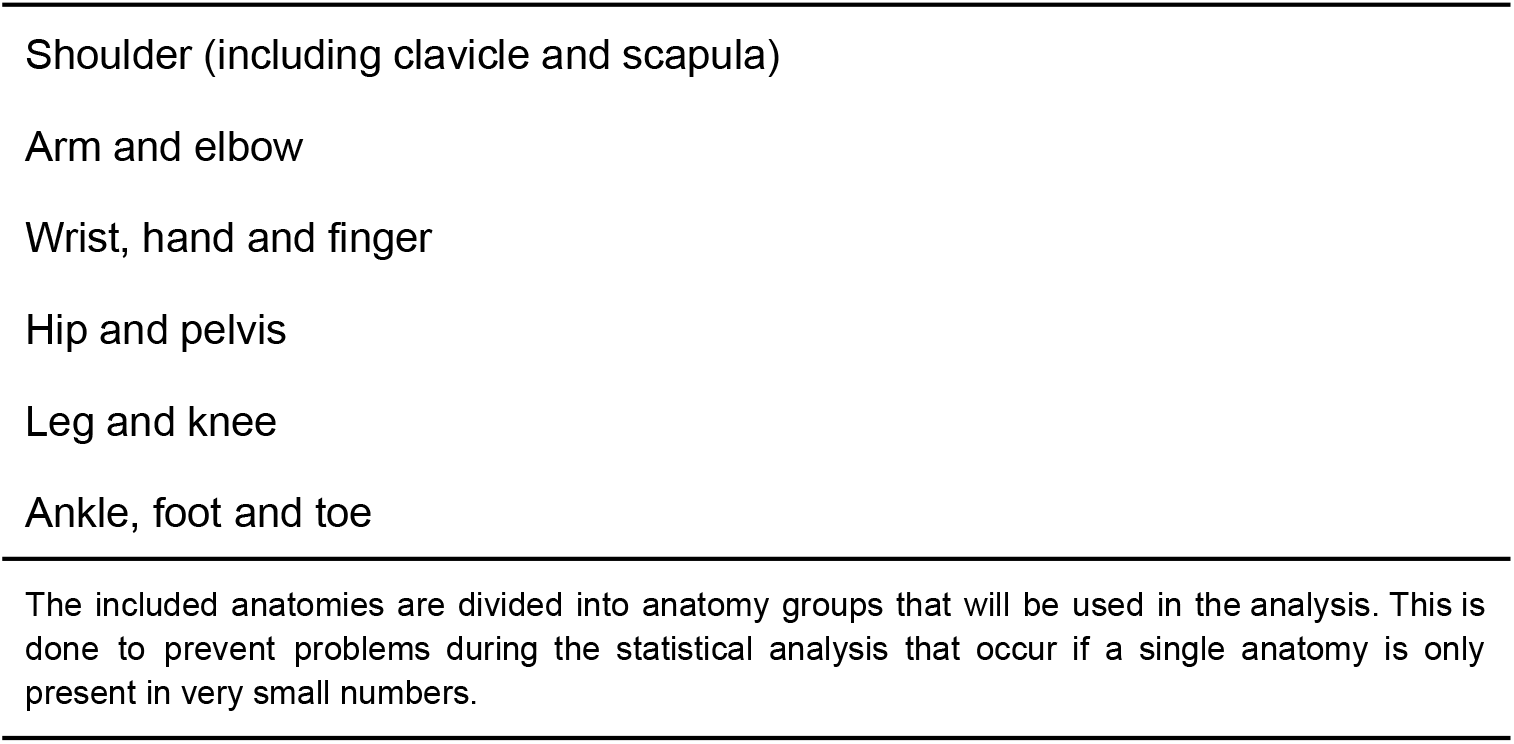
Anatomical subgroups.

### Data management

Imaging data and supplementary information as described in Table 1 are collected, anonymized and stored in a dataset locally at each site. This dataset will then be made available to the study participants via the annotations platform.

The generated results will be stored in the cloud and accessible to the relevant stakeholders across the sites. This dataset lacks both written and imaging data and does not contain sensitive data. No additional security measures are needed.

### Statistical methods

#### Input processing

Confidence ratings will be dichotomized by treating “definitely fractured” and “likely fractured, needs more imaging” as a detected fracture, and treating “likely not fractured, needs more imaging” and “definitely not fractured” as no fracture detected. Dots that fall within a standard reference bounding box are registered as detected fractures (Figure 6).

**Figure 5:**
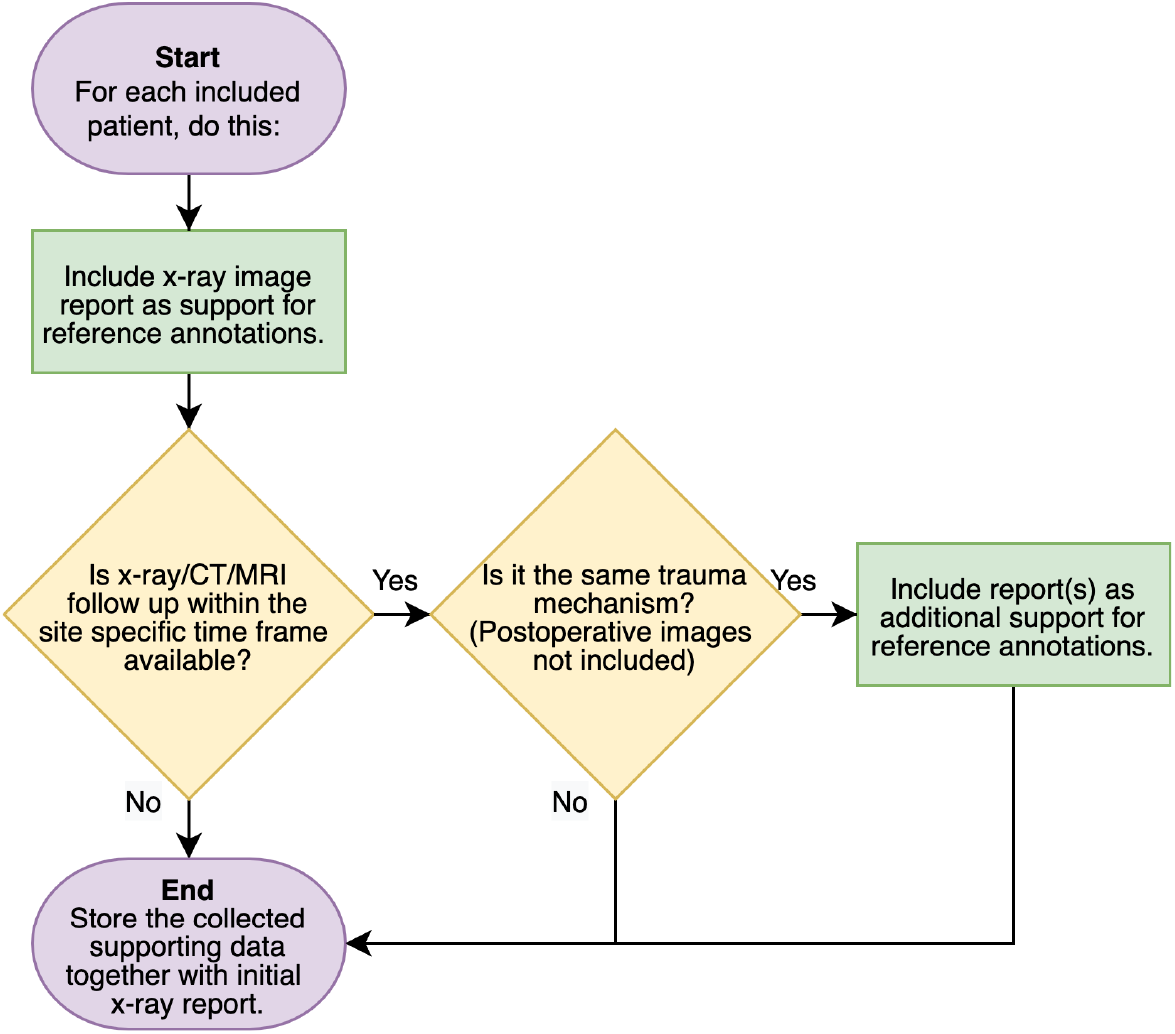
The above chart will be used to collect data of fracture status and the location of fracture. Additional imaging and expert opinion are assessed to find a conclusion.

**Figure 6.**
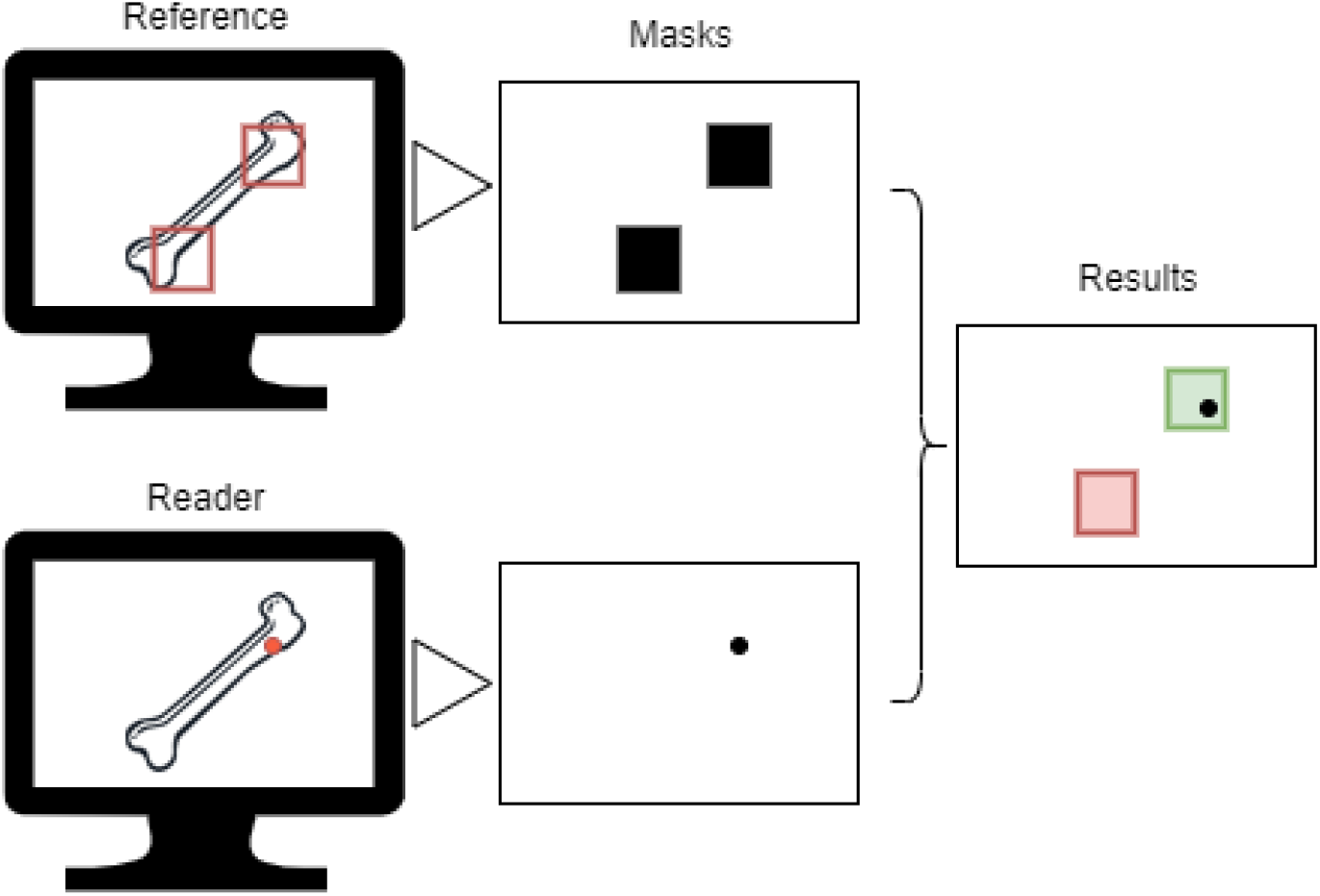
Dots that fall within a standard reference bounding box are registered as detected fractures. In this example, the reader has correctly identified the first fracture (green), while the second fracture was missed (red).

#### Metrics

The analysis will be based on the following metrics:

- The patient-wise sensitivity (SE_PW_), defined as the proportion of patients in whom all fractures are detected (each unique fracture in at least one radiograph). Note that this metric is not influenced by any potential incorrect marks (false positives).
- The patient-wise specificity (SPE_PW_), defined as the proportion of patients in whom no fracture mark was detected amongst patients without any fracture.
- The fracture-wise sensitivity (SE_FW_), defined as the proportion of fractures correctly identified by the reader amongst all fractures, counting multiple fractures per patient where appropriate.
- The average number of false-positive fractures per patient (FP_PPFW_), defined as the total number of marks put outside of a fracture divided by the number of patients.

#### Analysis

The primary endpoint is the impact on readers’ performance without and with AI-aid. This will be assessed by measuring the change in observed sensitivity and specificity (ΔSE_PW_, ΔSPE_PW_) on the patient-wise level; and the sensitivity and average number of false-positive fractures per patient (ΔSE_FW_, ΔFP_PPFW_) on the fracture-wise level using the definitions provided above.

The patient-wise metrics will be complemented by summary (SROC) ROC curves and corresponding AUCs will be calculated using the approach described by Oaken-Rayner et al.^7^ This approach, previously known from meta-analysis, treats each reader as a distinct diagnostic study and hereby prevents underestimation of human performance. For the fracture-wise level, free-response ROC curves (FROC) and corresponding AUCs will be calculated.

The metrics will be performed for each reader and the effect on performance will be compared between the three different sites. The stand-alone AI performance will be assessed using the same methods.

The success criteria of the primary analysis is defined as superiority of sensitivity and noninferiority of specificity with margin 5%. Paired t-tests will be used to compare by means of paired Student’s t-tests with one pair of observations for each reader. 95% confidence intervals for the metrics will be calculated using normal approximation to underlying data distribution. Normality assumption will be tested by Sharipo’s test and p-values of <0.05 are considered statistically significant.

#### Subgroup analysis

Patient-wise sensitivity and specificity, as well as fracture-wise sensitivity and false positive rate will be assessed for the individual anatomy groups. A minimum required number of positive fractures for each anatomy subgroup is set to n=20. Any group not meeting this requirement will be excluded from the subgroup analysis.

A second subgroup analysis will be performed in patients with multiple fractures. Performance gain of all readers and individual AI performance are recalculated for patients that have more than one fracture defined in the reference standard.

## Ethics and dissemination

### Research ethics approval

The study has received approval from the local research ethics committee (REC) or institutional review board (IRB) at each participating study site who waived the requirement of collecting informed consent prior to inclusion of retrospective data.

### Protocol amendments

Important protocol modifications (eg changes to eligibility criteria, outcomes and analyses) must be communicated to all investigators and, if necessary, communicated to the respective national ethical authorities.

### Confidentiality

Ethical considerations regarding the confidentiality of personal information about participants have been taken into account as much as possible. All data included in the study is anonymized and the results of the reading sessions that are used in the overall analysis do not contain information that enables assigning an identity to included patients. There is no physical intervention that can cause potential harm to the patient. During the reading sessions, clinical data from the referral that may have influence on reader interpretation should be kept available. Clinical information from the report that is not described in the table above is removed to avoid traceability. All data that issues privacy concerns and is not used to produce study results will be removed from the images such as name and patient ID.

### Access to data

All investigators and the sponsor will have access to the final, anonymised study dataset as recorded in the database.

### Dissemination policy

The findings from this study will be disseminated through publications in scientific as well as presentations at national and international conferences. Authorship will be given according to the guideline for authorship published by the International Committee of Medical Journal Editors.

## Data Availability

All data produced in the present study are available upon reasonable request to the authors.

## Abbreviations

AI: Artificial Intelligence
AUC: Area under the receiver operating characteristics curve
BFH: Bispebjerg and Frederiksberg Hospital
CUB: Charité - Universitätsmedizin Berlin
ED: Emergency Department
EMC: Erasmus Medical Center
IRB: Institutional Review Board
PACS: Picture Archiving and Communication System
REC: Research Ethics Committee

